# Impact Of Temperature and Sunshine Duration on Daily New Cases and Death due to COVID-19

**DOI:** 10.1101/2020.06.13.20130138

**Authors:** Swati Thangariyal, Aayushi Rastogi, Arvind Tomar, Ajeet Bhadoria, Sukriti Baweja

**Author notes:** **Corresponding Author:** Dr. Sukriti Baweja, PhD. **These authors contributed equally to this work.** (Swati Thangariyal, Aayushi Rastogi). **Financial Support:** The author(s) received no specific funding for this work. **Author’s Contribution: Swati Thangriyal:** Data collection, Methodology, Visualisation and Writing. **Aayushi Rastogi:** Data curation, Data analysis, Methodology, Statistics, Writing. **Arvind Tomar:** Conceptualisation, Writing - review and editing. **Sukriti Baweja:** Conceptualisation, Supervision, Methodology, Writing, Review and editing.

## Abstract

**Background:** The coronavirus pandemic (COVID-19) control has now become a critical issue for public health. Many ecological factors are proven to influence the transmission and survival of the virus. In this study, we aim to determine the association of different climate factors with the spread and mortality due to COVID-19.

**Methods:** The climate indicators included in the study were duration of sunshine, average minimum temperature and average maximum temperature, with cumulative confirmed cases, deceased and recovered cases. The data was performed for 138 different countries of the world, between January 2020 to May 2020. Both univariate and multivariate was performed for cumulative and month-wise analysis using SPSS software.

**Results:** The average maximum temperature, and sunshine duration was significantly associated with COVID-19 confirmed cases, deceased and recovered. For every one degree increase in mean average temperature, the confirmed, deceased and recovered cases decreased by 2047(p=0.03), 157(p=0.016), 743 (p=0.005) individuals. The association remained significant even after adjusting for environmental such as sunshine duration as well as non-environmental variables. Average sunshine duration was inveserly correlated with increase in daily new cases (ρ= -2261) and deaths (ρ= -0.2985).

**Conclusion:** Higher average temperature and longer sunshine duration was strongly associated with COVID-19 cases and deaths in 138 countries. Hence the temperature is an important factor in SARS CoV-2 survival and this study will help in formulating better preventive measures to combat COVID-19 based on their climatic conditions.

## Introduction

In December 2019, a novel virus with pneumonia-like illness was reported in a cluster of patients at the Central Hospital of Wuhan, China^1^. The new member of the Coronavirus family known as ‘SARS-CoV-2 (2019) with unknown etiology increased rapidly resulting in an outbreak associated with seafood markets in the city of Wuhan^2^. Since then it has spread almost all over the globe infecting ∼ 4.5M people in more than 200 countries and territories^3^. The rate of infection is very rapid that first it was declared as a public health emergency of international concern on January 31, 2020, and later its status was upgraded to pandemic. Climate conditions such as temperature, humidity, wind speed, solar radiation exposure, population density, and medical care quality are many factors that affect the transmission of the virus^4^. For the control and prevention of SARS-CoV-2 pandemic, the understanding of the characteristics of a geographical region with the spreading of COVID-19 is important. We assume that the climate conditions might also contribute to the infectivity of COVID-19. The United States of America and the European Region have reported the maximum number of confirmed cases and deaths related to COVID-19, whereas the African region has reported the minimum number of confirmed cases and deaths^3^. It is a noticeable fact that the WHO-South-East Asian region (SEAR) comprises approximately 23% of the world’s population but accounts for only 2.9% of the COVID-19 cases worldwide, However, second most populated country such as India is at the seventh position in the WHO list ofconfirmed cases with mortality rates between 2.5-2.9%^5^.

In the case of the Influenza virus study, the survival and transmission rate shows significant correlations with absolute humidity^6^. With the increase in the warm weather, a gradual decrease was seen in the 2003 outbreak of severe acute respiratory syndrome(SARS) cases in Guangdong and complete resolve at the end of July 2003^7^. Absolute humidity affects the seasonal outbreak of influenza according to the epidemiological model conducted in the United States^8^. Notably in the current pandemic of COVID-19, the role of environmental factors play an important role as suggested by data of epidemiology laboratory^9^. Change in climate conditions governs the Spatio-temporal spread of the virus^10^. Studies from Spain and Finland show 95% of infection worldwide falls in a dry region with temperature between ∼2°C -10^°^°C^11^. The association between temperature and rate of infection is also seen in the case of COVID-19. With the mean temperature below 3°C, there is an increase in 4.861 daily confirmed case ^12^. A positive association and negative association is reported by Ma et al. in March 2020^13^ between daily death by COVID-19, diurnal temperature, and relative humidity respectively but that only from Wuhan, China. In the warmer region, the spreading of COVID -19 was less in China indicated by showing the correlation between humidity, temperature, and COVID-19 outbreak^4^. Human movement, age, gender, comorbidities of individuals and ability to socialise play an important role in transmission of the virus^14^. The SARS-CoV-2 virus has shown a better stability at low temperature and low humidity^15^.Another study suggested exposure of SARS-COV to 56°C temperature for 90 minutes can deactivate the virus indicating survival difficulty of the virus at higher temperatures^16^. However, many other contradictory studies are reporting no association between the meteorological conditions and spread of COVID-19^17^,^18^. There is uncertainity in the association between the environmental factors and transmission susceptibility of the virus. Till date, there is no study that has studied the association between the sunshine duration and temperature from such large cohort of population, as a proof of concept. Hence, our aim of the present study, was to assess the effect of temperature, sunshine duration on confirmed, recovered and deceased cases between January 2020 to May 2020. There is an urgent need for better planning, regulation for preventive measures to combat the COVID-19 pandemic and our study will pave way for better understanding and may help the government in formulating better preventive measures based on climatic conditions of the region.

## Methods

### Data Collection

In silico, we collected the data of confirmed, recovered, and death cases of COVID-19 from an open-source compiled by the Johns Hopkins University Center for Systems Science and Engineering (JHU CCSE) https://data.humdata.org/dataset from 22^nd^ January 2020 till 17^th^ May 2020^19^ from all the countries, taking into account the range of countries with maximum - minimum average daily new cases and death in COVID-19. The data related to the maximum and minimum temperature for all the countries were retrieved from Accuweather online for the five months from January 2020 to May 2020^20^. However, it was difficult to gather information on all the locations in a particular country, we restricted ourselves to the capital cities of each country under consideration. Hence, we collected the data on maximum and minimum temperature and sunshine duration information only for the capital city and considered it to be representative of the entire country. The various variables that can be the potential confounders in the association between temperature and transmission of COVID-19 like Population, the median age of the country, Global Health Security (GHS) Rank, were also considered in the analysis. The data related to population and the median age was downloaded from https://ourworldindata.org/grapher^21^. GHS score and GHS rank from Global Health Security Index-2019 was studied as a proxy for healthcare preparedness of each country^22^. Since the main aim of our study was to assess the association between transmission and cause of COVID-19 death and temperature or sunshine duration, we assumed that countries having more than 300 cases are either having a cluster of cases or community transmission and hence excluded countries having less than 300 cases. We also excluded Diamond Princess from the analysis as it was a cruise ship and not a country as a whole.

### Data Analysis

All the data were analyzed using the SPSS software version 22 software (IBM SPSS). The Spearman’s correlation analysis was used to assess the association between temperature and confirmed, recovered and death cases of COVID-19. Followed by correlation analysis, we performed univariate and multivariate analyses. We assumed that the exposure and the outcome variable are following normal distribution and are having a linear relationship for simplicity. We checked the other assumptions for linear regression and failing to fulfill them, we used a robust option for further analysis. We prepared various multivariate models for the analysis to understand the association better. The first model included another environmental variable: Sun-shine duration. In the second model, we included other non-environmental variables which potentially can affect the association like GHS score, the median age of the population, total population per 1,00,000 individuals.

## Results

The data retrieved from Jan 22, 2020 to May 17, 2020, reported a total of 45, 25, 497 confirmed positive cases of COVID-19 infection spread over 213 countries in the world. Table 1 represents the global burden of confirmed, recovered, and death due to COVID-19 in various WHO-Regions till May 17^th,^ 2020^23^ showing maximum deaths in Region of America (AMRO) and minimum deaths in African region(AFRO).

**Table 1:** Global burden of COVID-19 in the WHO-regions.

| WHO-Regions | Confirmed cases | Deaths | Recovered Cases |
| --- | --- | --- | --- |
| African Region (AFRO) | 58663 | 1710 | 23480 |
| Region of the Americas (AMRO) | 1966932 | 118799 | 512810 |
| Eastern Mediterranean Region (EMRO) | 335088 | 9916 | 165594 |
| European Region (EURO) | 1870545 | 165951 | 855847 |
| South-East Asia Region (SEARO) | 135036 | 4411 | 48911 |
| Western Pacific Region (WPRO) | 167755 | 6737 | 145523 |

However the Figure 1 shows the global cumlulative trend in cases of COVID 19 from January 2020 to May 2020, clearly showing larger exponential growth of number of cases than recovered cases globally.

**Figure 1:**
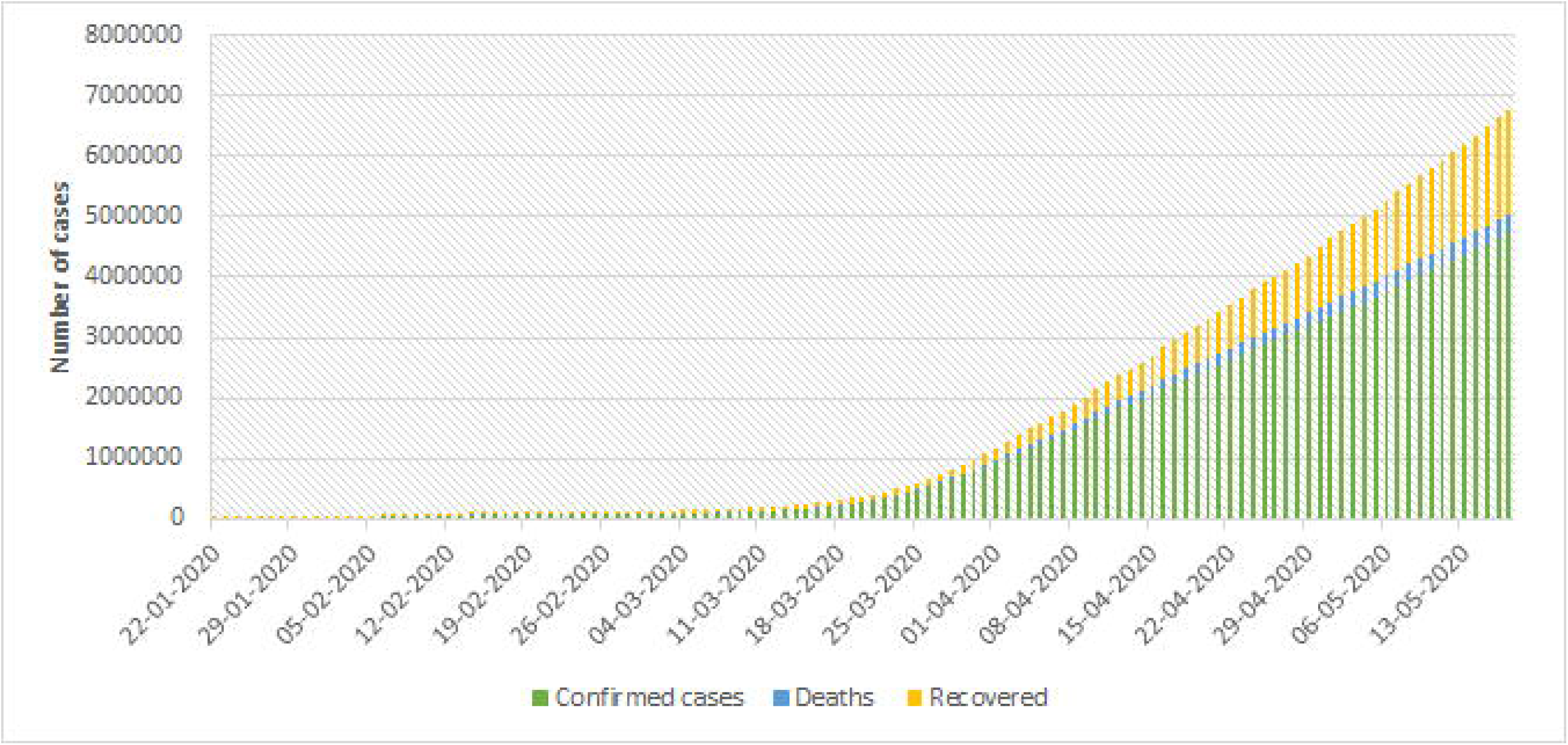
Global cumulative increase in number of new cases, death due to SARS-CoV-2 from all the countries affected from January 2020 to May 2020.

**Figure 2:**
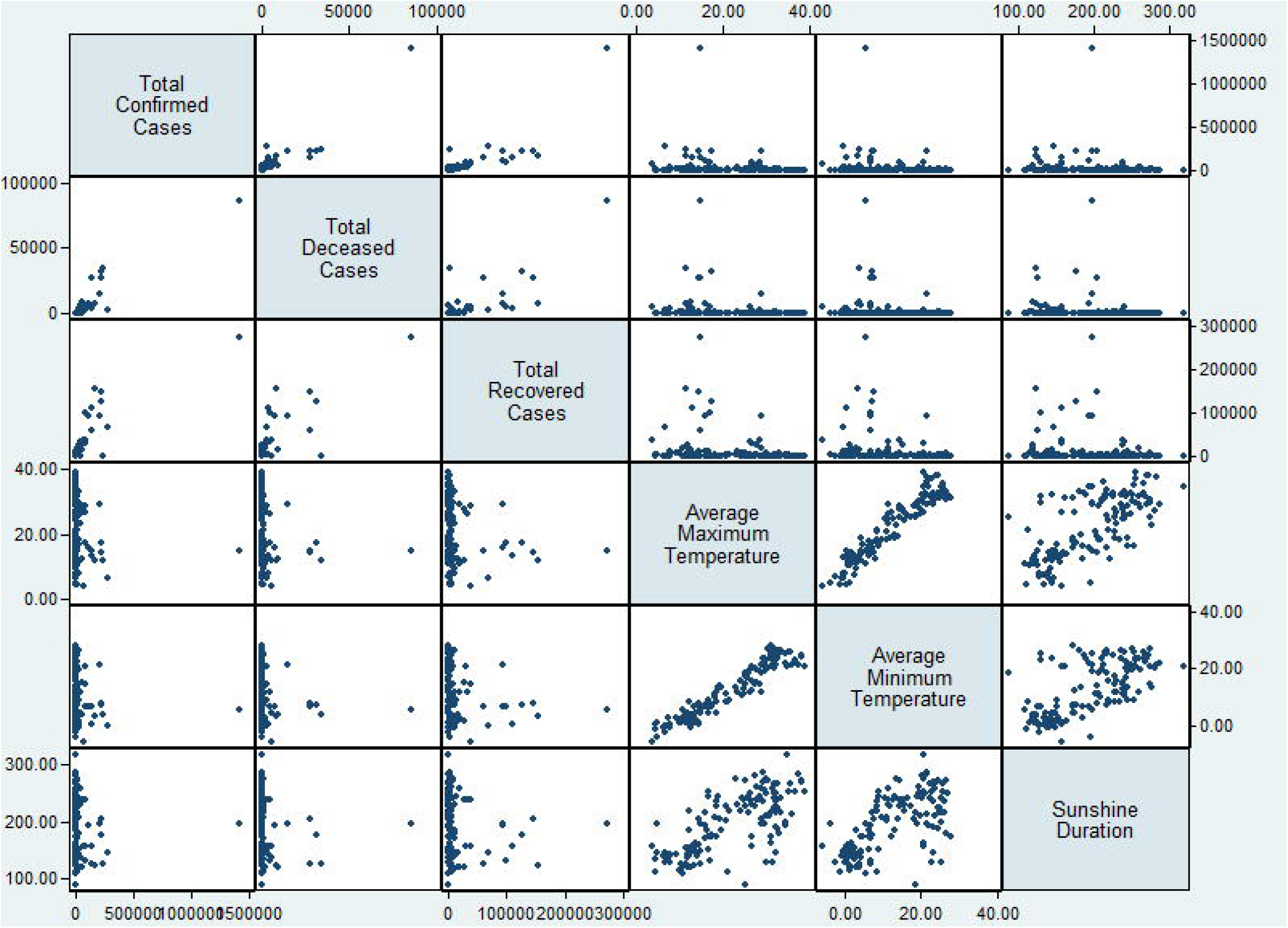
Scatter plots for each variable of minimum, maximum average temperature and duration of sunshine to determine the correlation between total confirmed cases, total deaths and total recovered cases.

Though the Wuhan, China was the origin of the infection, at present, the ten countries - United States of America (USA), Russian Federation, United Kingdom, Spain, Italy, Brazil, Germany, Turkey, France, and Iran account for more than 70% of the burden of the confirmed cases. Moreover, the majority of COVID-19 related deaths are majorly contributed by the USA, Italy, France, The United Kingdom and Spain whereas the maximum recovery have been observed in countries like the USA, Germany, Spain, Italy, and Turkey. These statistics indicate countries that are highly under impact of COVID-19 cases and mortality are also showing a speedy recovery. Further, these could be an influence due to environmental factors like temperature and sunshine duration.

### Correlational Analysis Between Mean Maximum Temperature And Cumulative Confirmed Cases, Death And Recovered Cases

We performed the anlaysis with 138 countries, having more than 300 confirmed cases of SARS-COV-2. The overall Spearman’s rank correlation analysis between average temperature (maximum and minimum) for five months and confirmed positive cases, dead and recovered cases of COVID-19 showed a weak and negative (−0.34) but significant association, indicating the severity of COVID-19 increases at low temperature. Further, a strong correlation (∼0.87) was observed between confirmed, recovered cases, and deaths for five months (Table 2). We also performed a similar correlation analysis for the five months separately (Supplementary 1). The number of confirmed cases represented a significant association with recovered and deceased cases each month separately, however, the correlation became stronger with an increased number of confirmed cases, i.e, from January 2020 to May 2020.

**Table 2:** Results of Spearman’s rank correlation

|  | Confirmed cases | Deaths | Recovered | Avg Max Temp | Avg Min Temp | Avg Sunshine duration |
| --- | --- | --- | --- | --- | --- | --- |
| Confirmed cases | 1.00 |  |  |  |  |  |
| Deaths | 0.8731* | 1.00 |  |  |  |  |
| Recovered | 0.8826* | 0.7728* | 1.00 |  |  |  |
| Avg Max Temp | -0.3406* | -0.3876* | -0.3544* | 1.00 |  |  |
| Avg Min Temp | -0.3374* | -0.4129* | -0.3675* | 0.9432* | 1.00 |  |
| Avg Sunshine duration | -0.2261* | -0.2985* | -0.2286* | 0.6891* | 0.6510* | 1.00 |
| * Indicates significant <0.05 |  |  |  |  |  |  |

### Univariate analysis between mean maximum temperature and Cumulative confirmed cases, Death and Recovered cases

Significant results were obtained on performing univariate analysis with temperature and confirmed cases, recovered cases, and death due to COVID-19 separately (Table 3). We also performed the same analysis for each month separately. The univariate analysis was not found to be significant for the month of January and February, whereas significant association (<0.05) with months of March and April. However, for the month of May data is marginally significant in recovered and death cases but not with confirmed cases (Supplementary II). A clear trend was visible, the increase in temperature, there is decline in number of cases anddeaths (Figure 3)

**Table 3:**
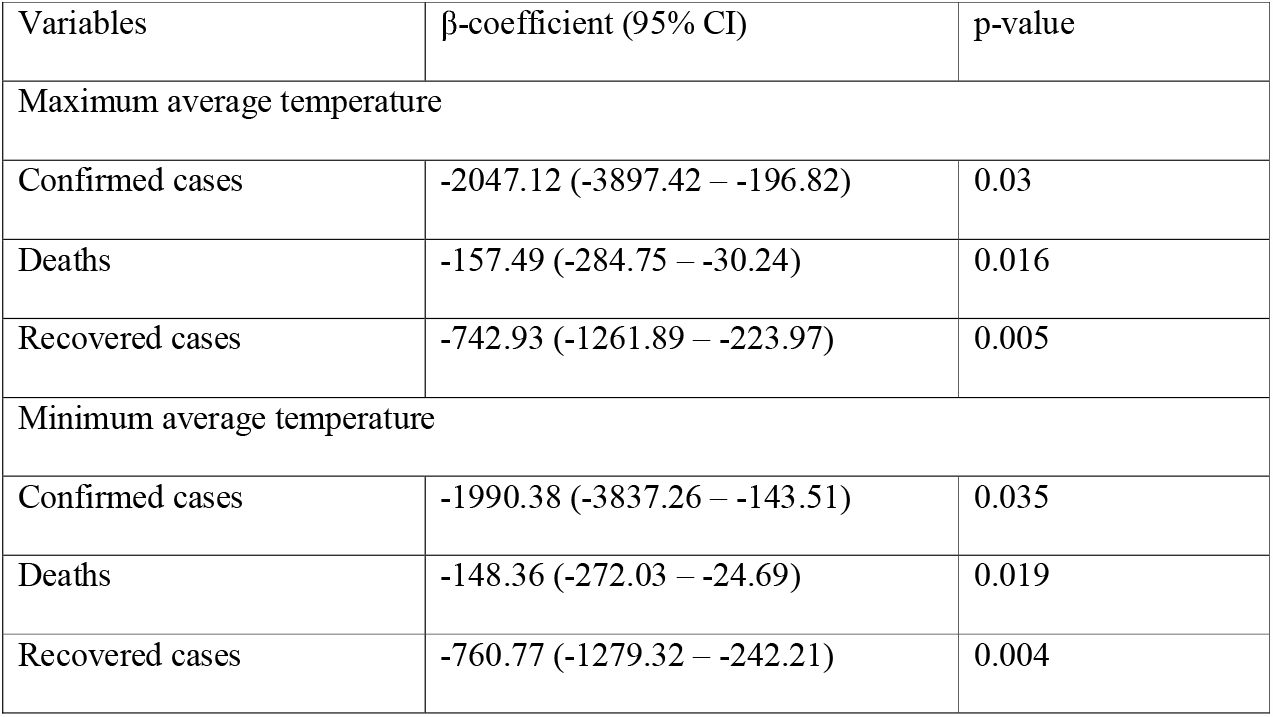
Univariate analysis of temperature with a confirmed case, recovered cases and death

| Variables | β-coefficient (95% CI) | p-value |
| --- | --- | --- |
| Maximum average temperature |  |  |
| Confirmed cases | -2047.12 (-3897.42 – -196.82) | 0.03 |
| Deaths | -157.49 (-284.75 – -30.24) | 0.016 |
| Recovered cases | -742.93 (-1261.89 – -223.97) | 0.005 |
| Minimum average temperature |  |  |
| Confirmed cases | -1990.38 (-3837.26 – -143.51) | 0.035 |
| Deaths | -148.36 (-272.03 – -24.69) | 0.019 |
| Recovered cases | -760.77 (-1279.32 – -242.21) | 0.004 |

**Figure 3:**
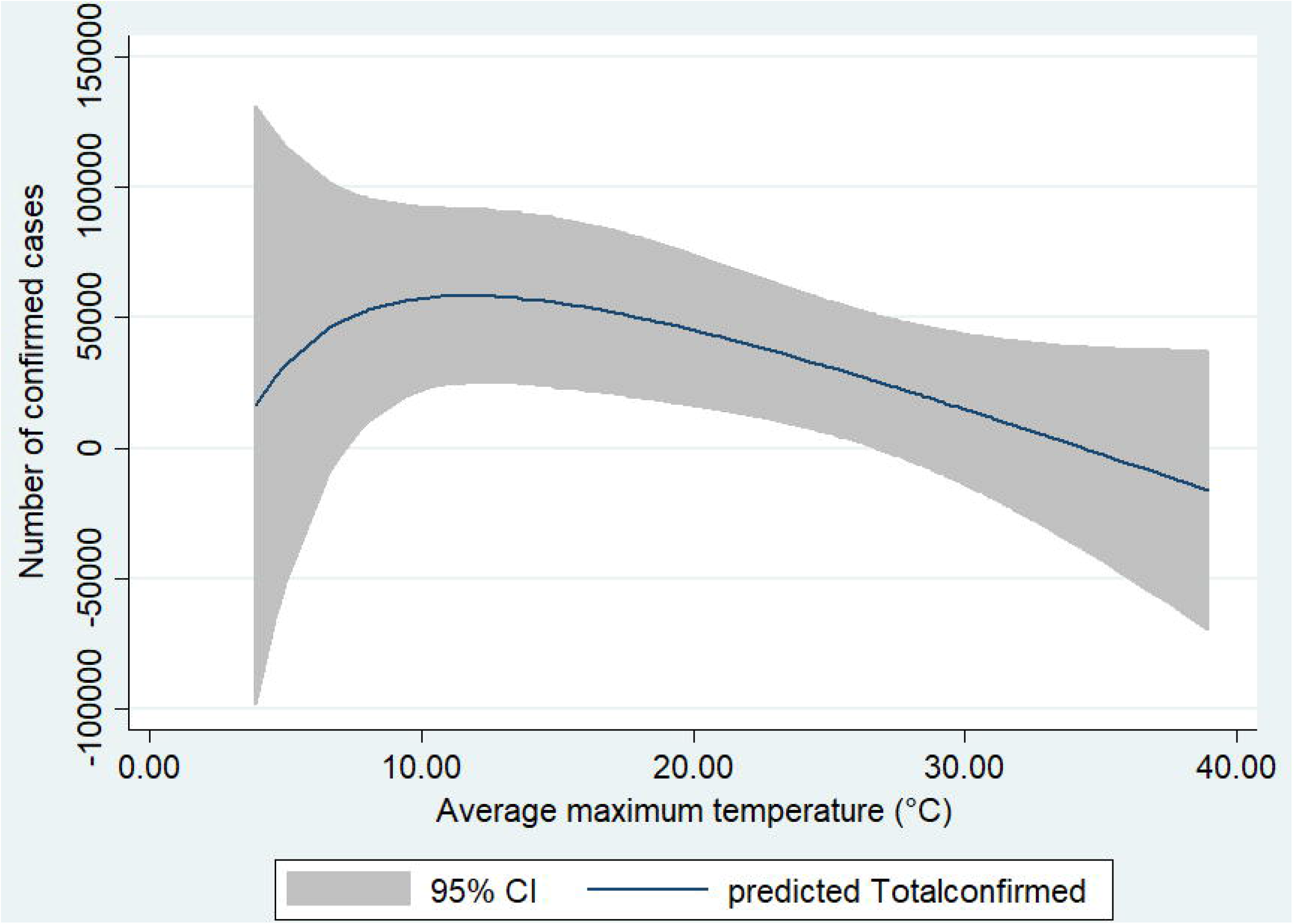
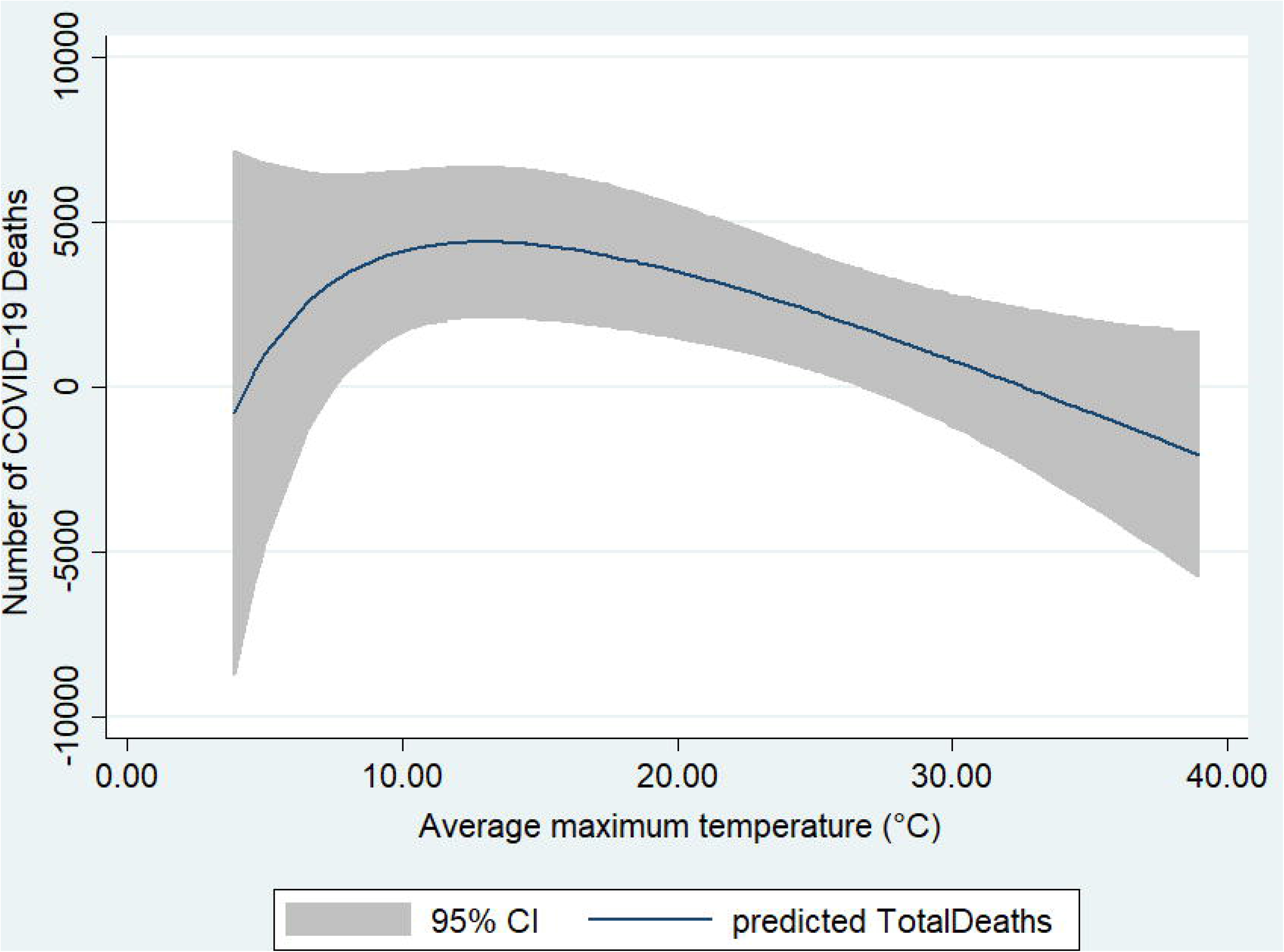
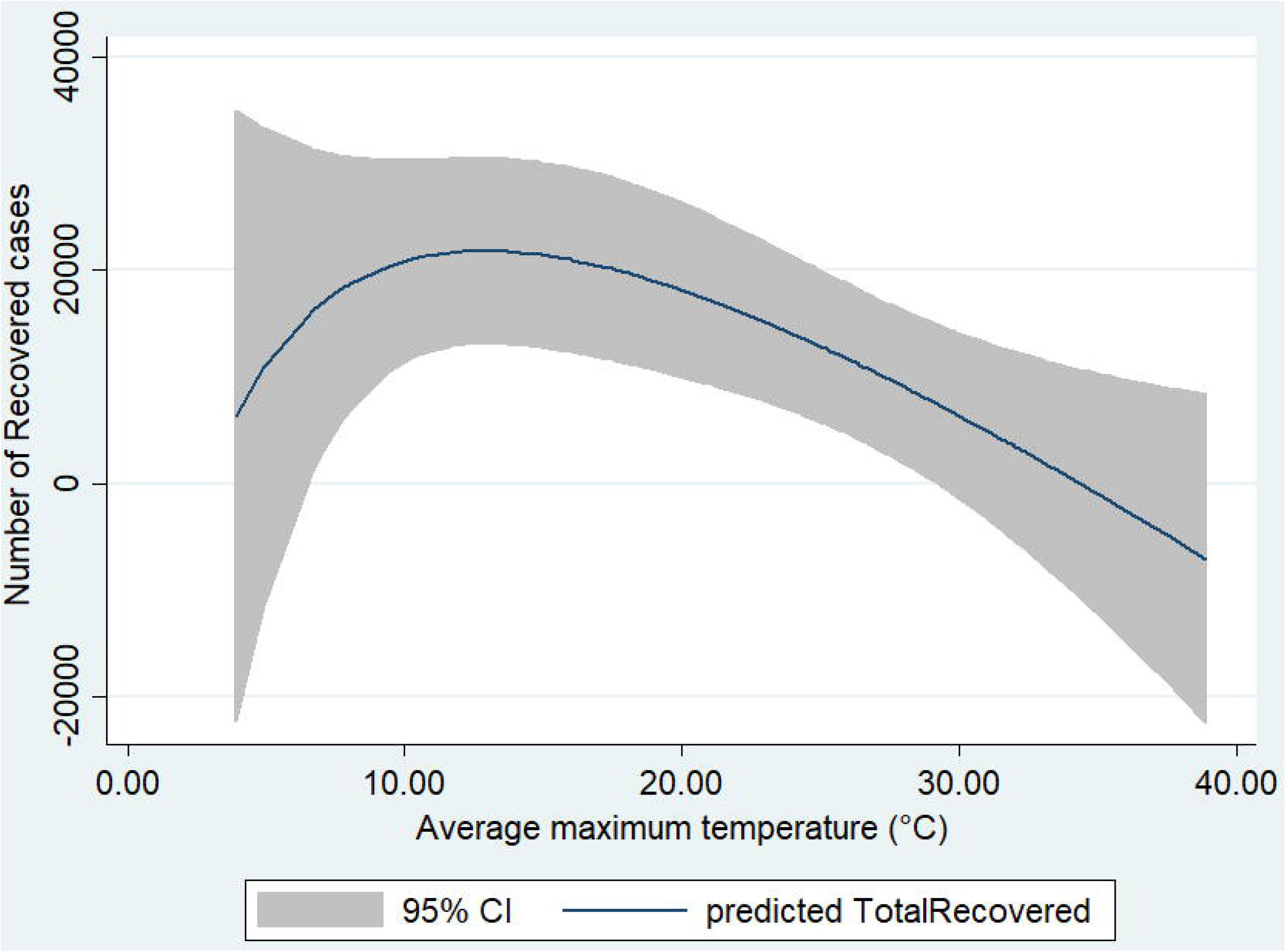
Association between Temperature and COVID-19 cases: (A) Association between temperature and confirmed cases. (B) Association between temperature and deceased cases. (C) Association between temperature and recovered cases.

### Multivariate regression analysis between mean maximum temperature and Cumulative confirmed cases, Death and Recovered cases

Further analysis with another environmental variable like sunshine duration showed a significant association between confirmed cases, recovered cases, death, and temperature (Table 4a).

**Table 4a:**
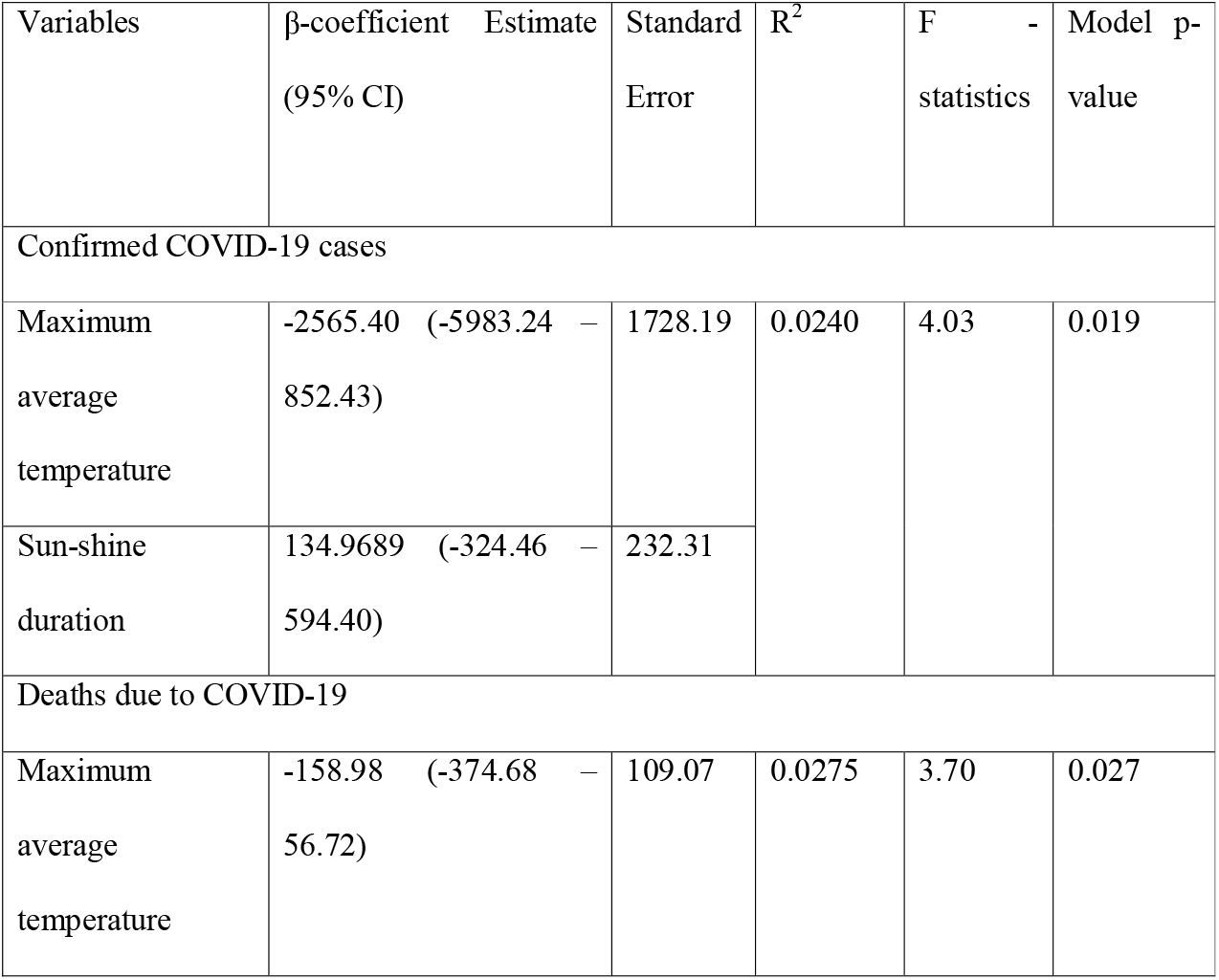
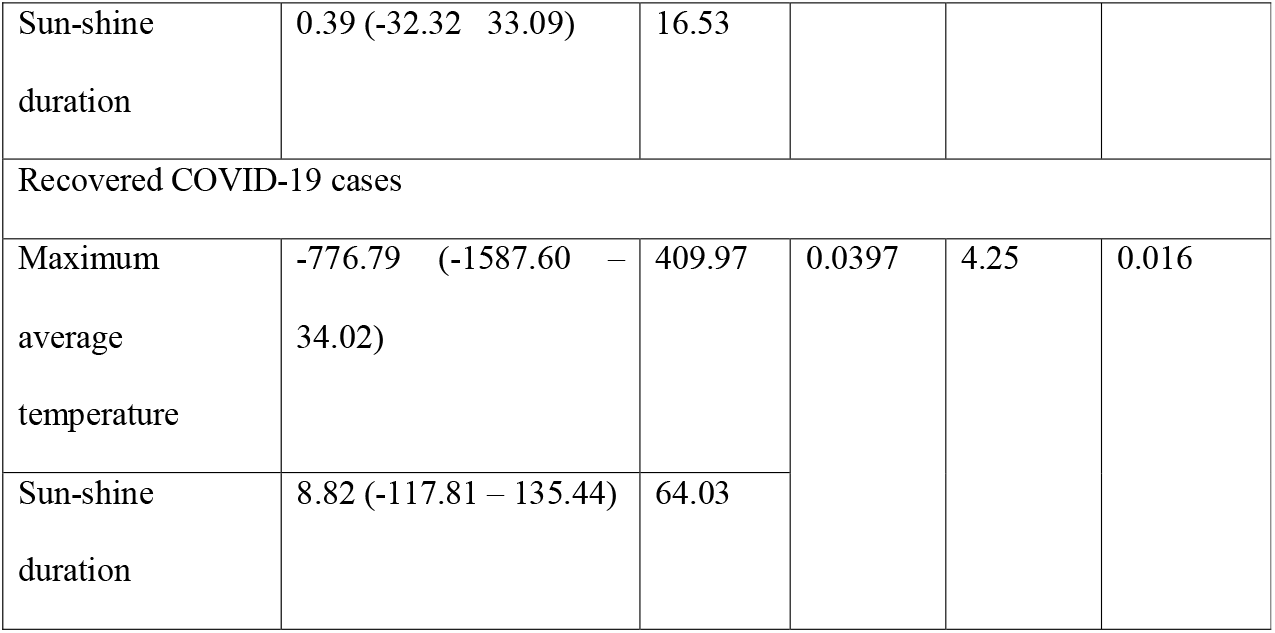
Adjusted analysis for association of COVID-19 and temperature

Table 4b suggests the association between temperature and confirmed cases, recovered cases and deaths remain significant (<0.05) even after adjusting for non-environmental factors like GHS score, GHS rank, the median age of the country, and population per 1,00,000 individuals. However, more factors can be added to make the model better.

**Table 4b:**
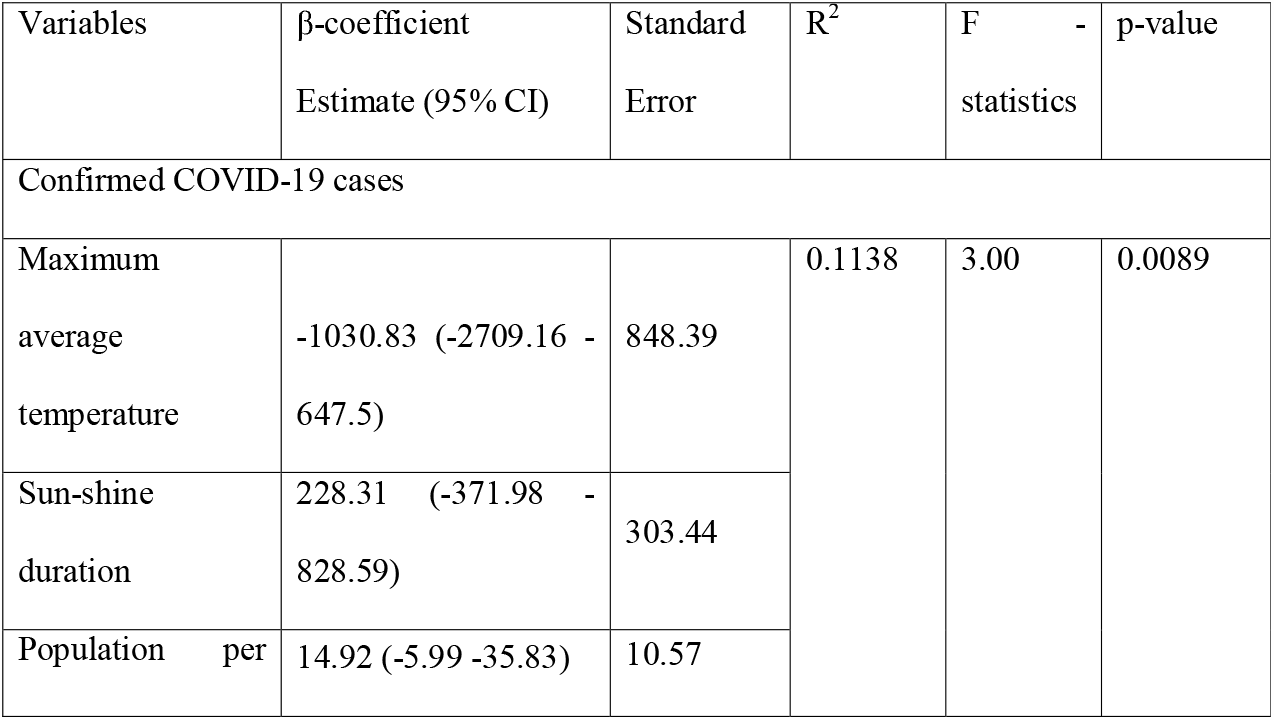
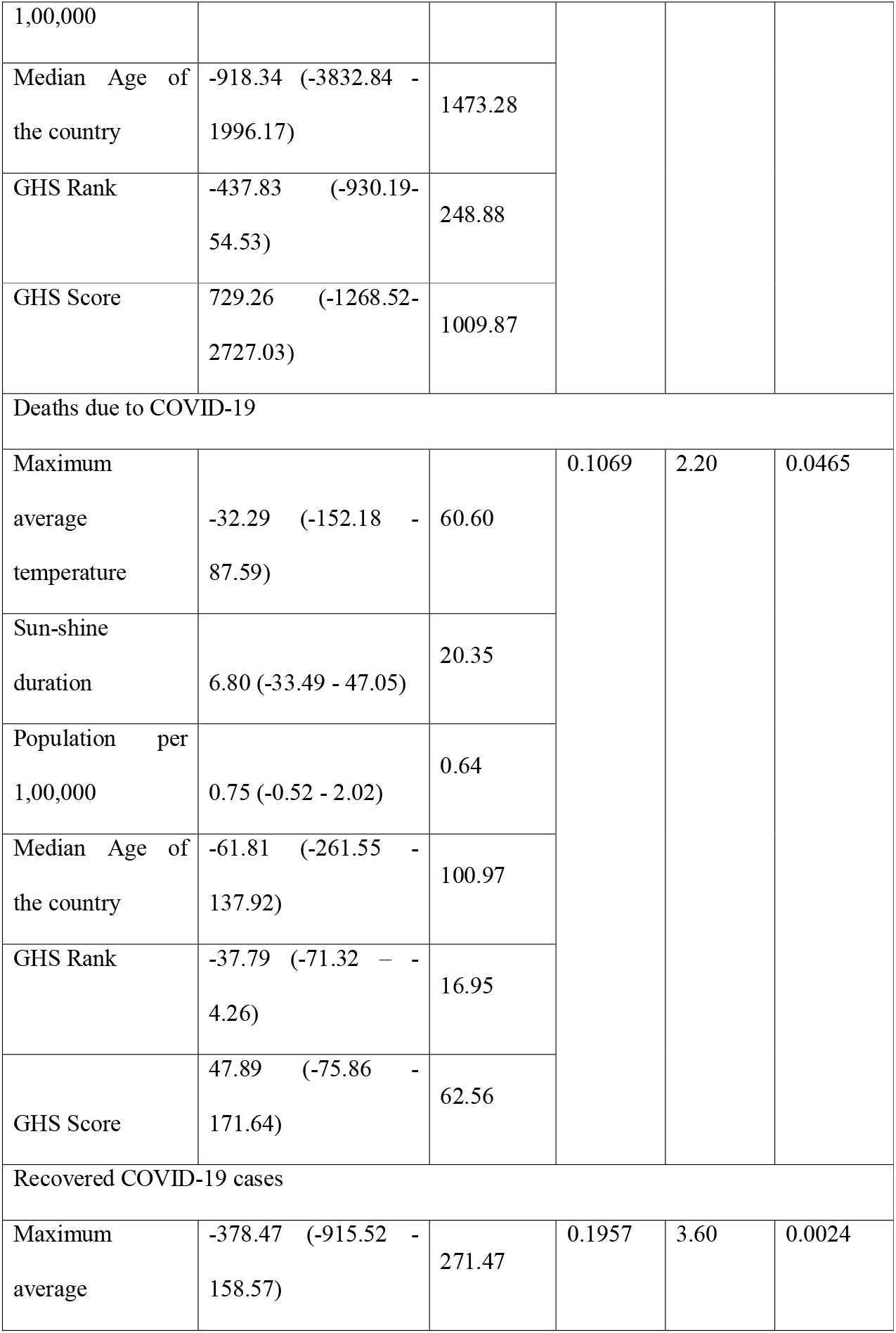
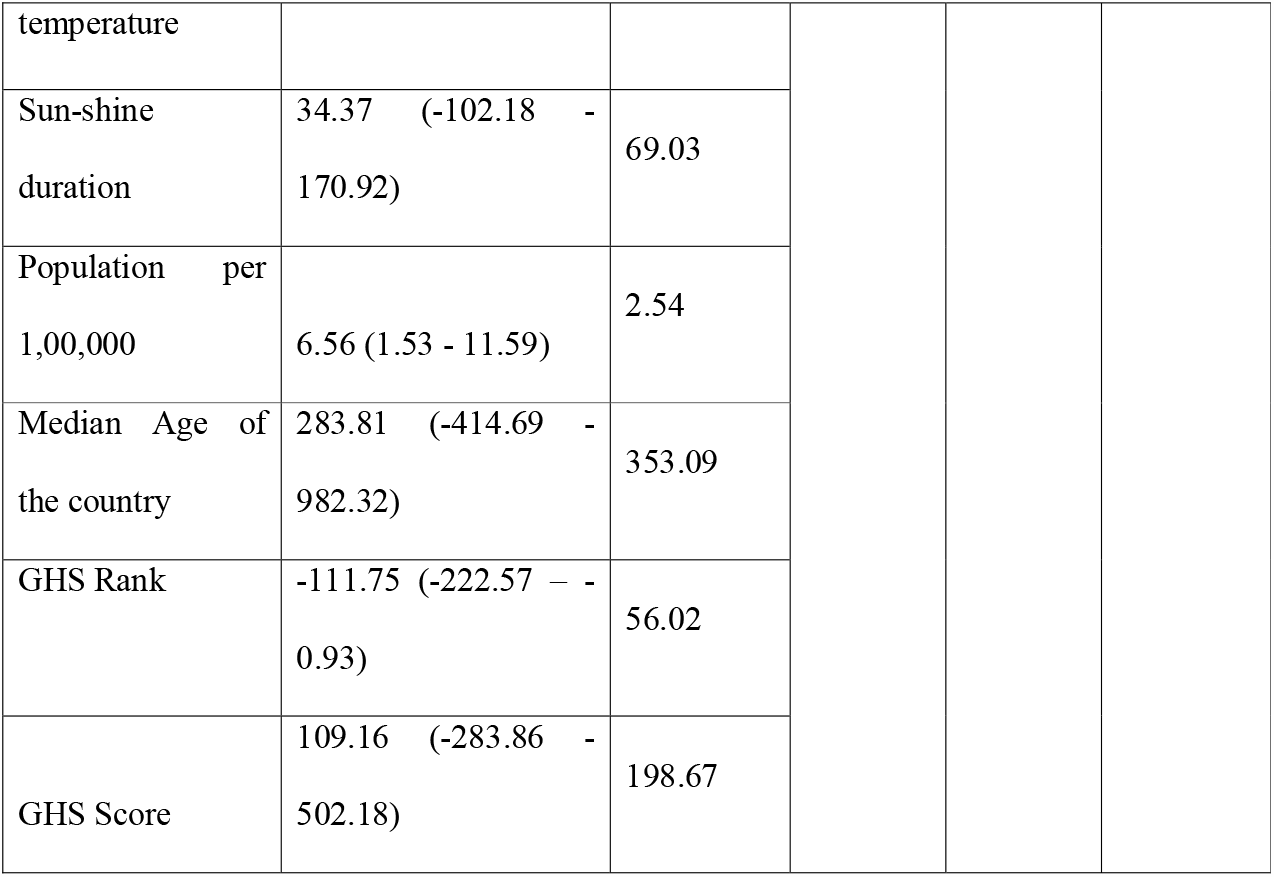
Adjusted Analysis for association of COVID-19 and temperature and other environmental and non-environmental variables

## Discussion

The spread and transmission of COVID-19 pandemic is a global emergency issue that is challenging for everyone. Our finding suggests the significant association between the temperature and sunshine duration with daily new cases and deaths of COVID-19. We anlaysed the data from January 22, 2020, to May 17, 2020, from 138 countries. Wuhan Province in China being the epicenter of the outbreak, occupies eighth place whereas American and Europen countries are at the top of the list. Most of the Asian countries are not available on the list of WHOhighly affected countries by novelcoronavirus. This unexpected distribution of data can be due to many reasons such as the genetics of the host, the immunization against different viral strains, mode of transmission of the virus, and climate conditions^24^.

This is a unique study which is addressing the association of temperature and sunshine duration with COVID-19 transmission after adjusting for non-environmental factors. The study also performed the cumulative as well as month-wise analysis.

In the correlation analysis confirmed cases, death, and recovered cases were found to be significantly correlated with average minimum,maximum temperature and sunshine duration (<0.05). Our study suggested a weak and negative correlation of temperature with the cumulative confirmed cases, cumulative deaths, and cumulative recoveries. Moreoverever, the average sunshine duration was also found to be significantly correlated with cumulative confirmed cases, cumulative deaths, and cumulative recoveries. Our results were found to be not coinciding with a similar study^25^ as the Yales study has taken data until the months of March whereas the present study has taken data till May 2020.

The univariate analysis suggested that the cumulative confirmed cases, recovered cases, and deaths due to COVID -19 were found to be significantly associated with mean maximum temperature and mean minimum temperature (<0.05). For every one-degree increase in mean average temperature, the confirmed cases, deaths, recovered cases decreased by 2047, 157, 743 individuals respectively. Though the results seems to be obvious, the decline in recovered cases can be attributable to the other factors such as an increasing burden on the health care system which we have not to addressed in our study.

After adjusting for environmental factors like sunshine duration, the association was found to be significant between mean maximum temp and cumulative confirmed cases, death, and recovered cases. However, the model had a weak R^2^ ranging from 2.40% to 3.97%. The association was still found to be significant with a p-value <0.05. After adjusting for non-environmental factors like GHS score, median age of the population, total population per 1,00,000 individuals, the R^2^ also improved and now ranges from 11.38% to 19.57%. A reduction of 1031 confirmed cases, 32 deaths, and 378 recovered cases were observed after adjusting for environmental and non-environmental factors. The decline in recovered cases can be attributable to other factors such as an increasing burden on the health care system.

Though our study is a unique study but there are several limitations in our study. The lacune of this study is that we assumed normal distribution of the outcome variable and linear relationship between exposure and outcome variable. However, the study was not able to address some kind of non-linearity in the analysis. Moreover, the data collected for the temperature and sunshine duration was only retrieved for the capital city and was assumed to be representative of the entire country and in reality it may not be the actual scenario. Since this study was retrospective, data for various variables was not available eg. humidity, airspeed, and most importantly the lockdown of human movement which is adopted by many countries. The total number of cases across the globe is difficult to predict due to the uncertainty of data collected by different countries. Hence, further evaluations are needed for a better understanding of the role of environmental and non environmental factors influencing the transmission of this pandemic. In conclusion, our study showed a possible association between environmental conditions and COVID-19 infection.

## Data Availability

The data is available and uploaded as supplementary material.

## Abbreviations

SARS: Severe acute respiratory syndrome
SARS-CoV-2: severe acute respiratory syndrome new Coronavirus2
GHS: Global Health Security
WHO: World Health Organisation
AFRO: African region
AMRO: Region of Americas
EMRO: Eastern Mediterranean Region
EURO: European Region
SEARO: South-East Asia Region
WPRO: Western Pacific Region

## Supporting Infromation

### Supplementary I

**Table 1:**
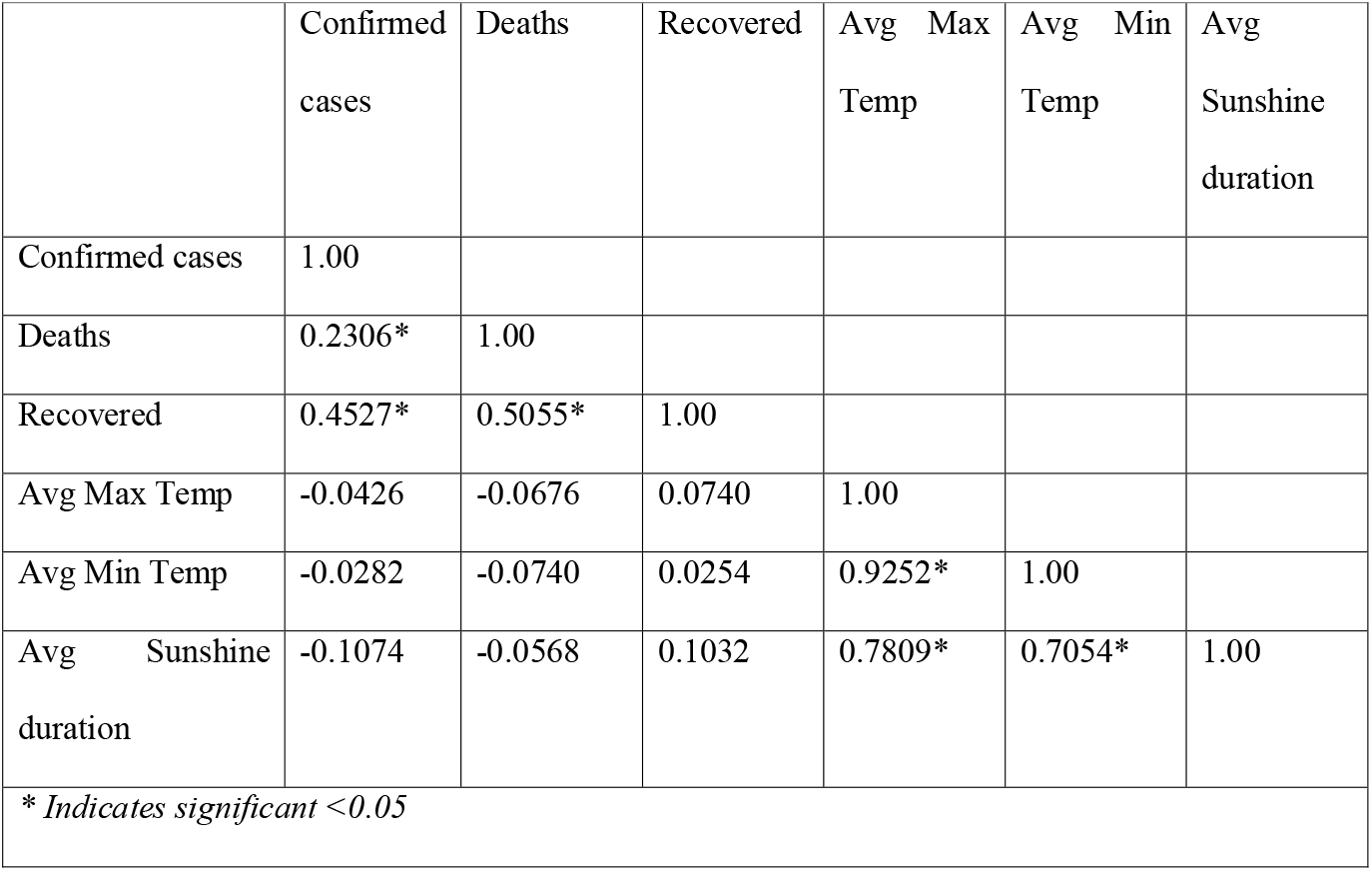
Results of Spearman’s rank correlation for the month of January, 2020

**Table 2:** Results of Spearman’s rank correlation for the month of February, 2020

|  | Confirmed cases | Deaths | Recovered | Avg Max Temp | Avg Min Temp | Avg Sunshine duration |
| --- | --- | --- | --- | --- | --- | --- |
| Confirmed cases | 1.00 |  |  |  |  |  |
| Deaths | 0.4075* | 1.00 |  |  |  |  |
| Recovered | 0.6437* | 0.5293* | 1.00 |  |  |  |
| Avg Max Temp | -0.2939* | -0.1050 | -0.1101 | 1.00 |  |  |
| Avg Min Temp | -0.2586* | -0.0996 | -0.0585 | 0.9264* | 1.00 |  |
| Avg Sunshine<br>duration | -0.2397* | -0.0985 | -0.0915 | 0.7822* | 0.7153* | 1.00 |
| * Indicates significant <0.05 |  |  |  |  |  |  |

**Table 3:** Results of Spearman’s rank correlation for the month of March, 2020

|  | Confirmed<br>cases | Deaths | Recovered | Avg Max<br>Temp | Avg Min<br>Temp | Avg<br>Sunshine<br>duration |
| --- | --- | --- | --- | --- | --- | --- |
| Confirmed cases | 1.00 |  |  |  |  |  |
| Deaths | 0.8551* | 1.00 |  |  |  |  |
| Recovered | 0.8184* | 0.6955* | 1.00 |  |  |  |
| Avg Max Temp | -0.5234* | -0.3818* | -0.3306* | 1.00 |  |  |
| Avg Min Temp | -0.5219* | -0.4010* | -0.3304* | 0.9482* | 1.00 |  |
| Avg Sunshine<br>duration | -0.3854* | -0.3085* | -0.2207 | 0.6983* | 0.6636* | 1.00 |
| * Indicates significant <0.05 |  |  |  |  |  |  |

**Table 4:** Results of Spearman’s rank correlation for the month of April, 2020

|  | Confirmed<br>cases | Deaths | Recovered | Avg Max<br>Temp | Avg Min<br>Temp | Avg<br>Sunshine<br>duration |
| --- | --- | --- | --- | --- | --- | --- |
| Confirmed cases | 1.00 |  |  |  |  |  |
| Deaths | 0.8760* | 1.00 |  |  |  |  |
| Recovered | 0.8347* | 0.7696* | 1.00 |  |  |  |
| Avg Max Temp | -0.3133* | -0.3992* | -0.3459* | 1.00 |  |  |
| Avg Min Temp | -0.3662* | -0.4737* | -0.4095* | 0.9212* | 1.00 |  |
| Avg Sunshine duration | -0.1506 | -0.2431* | -0.1717* | 0.5192* | 0.5136* | 1.00 |
| * Indicates significant <0.05 |  |  |  |  |  |  |

**Table 5:** Results of Spearman’s rank correlation for the month of May#, 2020

|  | Confirmed cases | Deaths | Recovered | Avg Max Temp | Avg Min Temp | Avg Sunshine duration |
| --- | --- | --- | --- | --- | --- | --- |
| Confirmed cases | 1.00 |  |  |  |  |  |
| Deaths | 0.8301* | 1.00 |  |  |  |  |
| Recovered | 0.7515* | 0.7041* | 1.00 |  |  |  |
| Avg Max Temp | -0.0170 | -0.2470* | -0.1841* | 1.00 |  |  |
| Avg Min Temp | -0.0723 | -0.3016* | -0.2437* | 0.9306* | 1.00 |  |
| Avg Sunshine duration | 0.0409 | -0.0364 | 0.0038 | 0.1771* | 0.1092 | 1.00 |
| * Indicates significant <0.05 |  |  |  |  |  |  |

### Supplementary II

**Table 1:** Univariate analysis of temperature with a confirmed case, recovered cases and death for the month of January 2020

| Variables | βcoefficient (95% CI) | p-value |
| --- | --- | --- |
| Maximum average temperature |  |  |
| Confirmed cases | -5.72 (-17.09 – 5.65) | 0.322 |
| Deaths | -0.13 (-0.37 – 0.12) | 0.321 |
| Recovered cases | -0.12 (-0.37 – 0.13) | 0.342 |
| Minimum average temperature |  |  |
| Confirmed cases | -6.07 (-18.16 – 6.01) | 0.322 |
| Deaths | - 0.13 (-0.39 – 0.13) | 0.321 |
| Recovered cases | -0.13 (-0.39 – 0.14) | 0.340 |

**Table 2:** Univariate analysis of temperature with confirmed case, recovered cases and death for the month of February, 2020

| Variables | β-coefficient (95% CI) | p-value |
| --- | --- | --- |
| Maximum average temperature |  |  |
| Confirmed cases | -32.63 (-91.29 – 26.02) | 0.273 |
| Deaths | -1.15 (-3.35 – 1.05) | 0.305 |
| Recovered cases | -16.40 (-49.19 – 16.39) | 0.325 |
| Minimum average temperature |  |  |
| Confirmed cases | -45.15 (-127.52 – 37.21) | 0.280 |
| Deaths | -1.61 (-4.71 – 1.48) | 0.306 |
| Recovered cases | -23.22 (-69.37 – 22.94) | 0.322 |

**Table 3:** Univariate analysis of temperature with confirmed case, recovered cases and death for the month of March, 2020

| Variables | βcoefficient (95% CI) | p-value |
| --- | --- | --- |
| Maximum average temperature |  |  |
| Confirmed cases | -341.32 (-556.12 – -126.53) | 0.002 |
| Deaths | -16.92676 (-31.44 – -2.42) | 0.023 |
| Recovered cases | -65.29 (-113.24 – -17.34) | 0.008 |
| Minimum average temperature |  |  |
| Confirmed cases | -338.60 ( -549.43 – -127.77) | 0.002 |
| Deaths | -15.79 (-29.45 – -2.13) | 0.024 |
| Recovered cases | -65.69 (-113.26 – -18.13) | 0.007 |

**Table 4:**
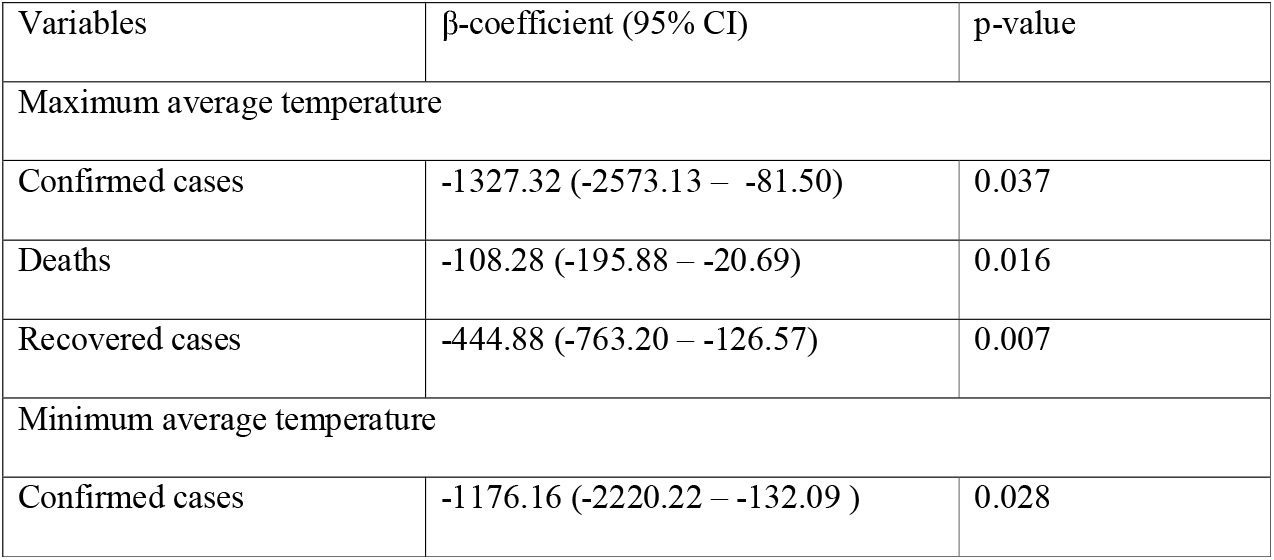
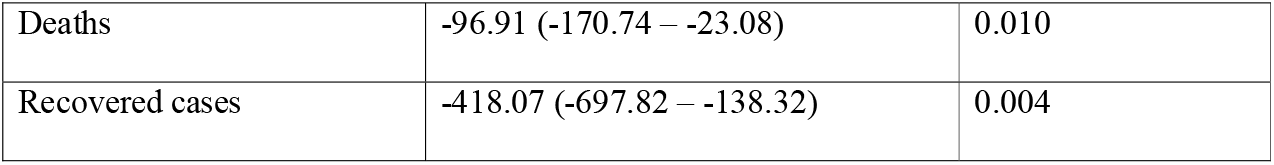
Univariate analysis of temperature with confirmed case, recovered cases and death for the month of April, 2020

**Table 5:** Univariate analysis of temperature with confirmed case, recovered cases and death for the month of May#, 2020

| Variables | βcoefficient (95% CI) | p-value |
| --- | --- | --- |
| Maximum average temperature |  |  |
| Confirmed cases | -499.97 (-1225.56 – 225.62) | 0.175 |
| Deaths | -51 (-103.28 – 1.27) | 0.056 |
| Recovered cases | -199.03 (-444.92 – 46.86) | 0.112 |
| Minimum average temperature |  |  |
| Confirmed cases | -491.52 (-1107.38 – 124.34) | 0.117 |
| Deaths | -46.35 (-91.74 – -0.96) | 0.045 |
| Recovered cases | -244.63 (-469.98 – -19.27) | 0.034 |

